# Translation, adaptation and psychometric evaluation of the German version of the Abortion Attitude Scale – a secondary analysis of a cross-sectional study among medical students

**DOI:** 10.1101/2025.03.14.25323947

**Authors:** Anja Lindig, Eva Christalle, Jördis Maria Zill, Mirja Baumgart

## Abstract

**Background:** Unintentionally pregnant individuals in Germany seeking an abortion face challenges due to legal regulations, stigma and difficult access to abortion care. Abortion attitudes of (prospective) physicians influence the care situation. To measure these attitudes, psychometrically sound instruments like the Abortion Attitude Scale (AAS) are necessary. So far, no instruments assessing attitudes toward abortions are available in German. The aim of this study is to translate, culturally adapt and psychometrically test the AAS.

**Methods:** This is a secondary analysis of a cross-sectional study on abortion attitudes of medical students in Germany. The English 14-item AAS was translated into German and adapted using a team translation protocol. Comprehensibility was tested via cognitive interviews (n=10 medical students). We analyzed acceptance (completion rate), factorial structure (confirmatory factor analysis (CFA), model fit), item characteristics (response distribution, item difficulties, corrected item-total correlations, inter-item correlations), and reliability (McDonald’s omega).

**Results:** The translated and adapted AAS version was comprehensible. AAS data of 305 medical students could be included in analysis. Completion rate was above 98% for all items. The CFA results confirmed a one-factorial structure but a model without item 10 and correlations between item 8 and item 13 showed to have the best model fits. Floor or ceiling effects could be found for 7 items, item difficulties ranged between 0.39 and 0.94, corrected item-total-correlations ranged between 0.460 and 0.766 for the best model, inter-item correlations ranged between .129 to .681, and McDonald’s omega was above 0.9 for both models.

**Conclusion:** The German AAS is a brief measure with high acceptance and good psychometric properties. Removal of item 10 could be discussed. The AAS can ease and improve the evaluation of attitudes toward abortions in Germany. This can potentially lead to the development of targeted interventions to reduce barriers and improve care for unintentionally pregnant individuals.

## INTRODUCTION

Attitudes towards abortion are a key construct for understanding societal trends in opinions, the impact of gender-based violence, and legislative changes surrounding reproductive rights in various countries (1). According to the Theory of the Triangle of Violence (2), negative attitudes towards abortion can manifest as gender-based violence across three interconnected dimensions: 1) *structurally*, restrictive abortion legislation in some regions constitutes a violation of human rights (3) 2) *culturally*, stigmatizing attitudes often stem from traditional and conservative values, such as prioritizing responsibility and caregiving over self-determination or reinforcing archetypes of femininity that equate womanhood with motherhood, 3) dire*ct violence*, which is expressed through individual actions and discriminatory treatment. These dimensions collectively contribute to abortion stigma, which extend beyond social dynamics by significantly impacting women’s mental health. Women who experience stigmatization report higher levels of depression, anxiety, stress, psychological distress, and somatic symptoms (4–6). The community’s abortion attitudes, therefore, have a direct bearing on women’s health and internalized stigma, underscoring the need for systematic exploration of these attitudes (1).

Conscientious objection to abortion adds another dimension to this topic. Defined as a physician’s refusal to perform legal abortions based on religious or moral beliefs, it is the only avenue for providers to refuse healthcare that would normally fall within their scope of practice (7). Conscientious objection is criticized for inadequately balancing abortion access with providers’ rights (7). It has been identified as a barrier to timely and safe abortion care, contributing to increased morbidity and mortality (8,9). Studies indicate that, depending on the country and its legal regulations, 14–80% of clinicians globally refuse to provide legal abortion services (8). Conscientious objection exists also in Germany (based on §12 SchKG, (10)) and leads to stigmatization of people seeking abortions and healthcare professionals working in abortion care (11–16) as well as limited access to abortion care (16,17). The Germany law allows abortion under certain conditions while maintaining its status as technically illegal under the German criminal code (§218 StGB, (18)): 1) within the first 14 weeks after conception and after undergoing mandatory counselling and a 3 day waiting period between counselling and abortion, 2) for medical reasons in case of expected psychological or physiological harm of the mother, and 3) for criminal reasons like sexual abuse. This regulatory framework created a nuanced and often polarized societal discourse (19–21). The lack of training on abortion procedures in most German medical school curricula reinforces the barriers for high quality abortion care in Germany (15,16). Thus, physicians often bear the responsibility of acquiring the necessary skills for abortion care independently. While the total number of abortions performed in Germany has remained stable (106.218 reported in 2023, (22)), the number of facilities offering abortion services (“Meldestellen”) has drastically declined over the past 20 years, dropping from 2.050 to 1.092 (23). A recent qualitative study involving 18 experts in psycho-social and medical abortion care highlighted that this shortage of providers significantly hampers the delivery of person-centered abortion care (16).

In this context, the need for psychometrically sound instruments to evaluate abortion attitudes becomes evident. Such tools are essential for addressing the complex interplay of stigma, conscientious objection, and reproductive rights to inform interventions or policies for future high quality abortion care, particularly within specific sociocultural contexts like Germany. There is especially a high need for evaluating attitudes of (future) physicians as a predictor for their (future) participation in abortion care (24). The theory of planned behavior postulates that attitudes (towards abortion), social norms and perceived control of action influence intentions (to perform abortions) and that perceived control of action predicts behavior (25). Accordingly, physicians having more positive attitudes towards abortion are more likely to provide abortions (26,27).

Existing self-report instruments on abortion attitudes, such as the Stigmatizing Attitudes, Beliefs, and Actions Scale (28) or the Abortion as a Right Scale (29) have been developed primarily in African countries like Ghana and Zambia, but the item content and focus of the scale does not align with the sociocultural realities of countries like Germany. Others like the Abortion-Providing Physicians Scale (30) or the Adolescent Attitudes to Abortion Scale (31) have a different focus by assessing attitudes of health professionals towards abortion providers or of adolescents. Furthermore, many instruments evaluate the level of agreement with abortion in a series of circumstances and not cognitions and beliefs (1,32). The scarcity of instruments with robust psychometric properties highlights a gap in tools designed for specific populations, such as those in Germany (1). Since there is no instrument on abortion attitudes in German language available, the translation of the Abortion Attitude Scale (AAS) into German is promising because the original has shown good reliability and construct validity, it is assessable for the German sociocultural background and it targets cognitions and beliefs (33). The AAS, developed by Linda A. Sloan in the United States in 1983, captures a range of perspectives on abortion, from personal beliefs to perception of societal norms and is usable in various settings (33,34). Alspaugh et al. (2021) used the AAS in the context of assessing attitudes of womens’ health and neonatal nurses in the U.S. but applied only slight modifications to reflect a more neutral, person-centered language without systematic adaptation or testing of the adapted items (35). Furthermore, they did not assess psychometric properties of the scale.

It is necessary to not only translate but also adapt an instrument to ensure that it maintains it’s conceptual and measurement equivalence in the target culture (36), since existing measures may not fully capture the cultural nuances and legal realities of other contexts. This includes potential differences in language semantics, cultural norms, and value systems that may influence how respondents interpret and respond to survey items (36). Furthermore, there is an increasing need for comparative research on abortion attitudes across countries. For such studies to be meaningful, the required tools have to be psychometrically robust across different settings and need to have similar characteristics.

Therefore, the aim of this study was to translate and culturally adapt the AAS instrument into German and evaluate its psychometric properties.

## METHODS

### Study design

We conducted this secondary analysis as part of a cross-sectional online study on influencing factors on abortion attitudes of medical students in Germany (24). Based on the Intergroup Contact Theory (37) and the Theory of Planned Behavior (25), the analyzed influencing factors included subjective and objective knowledge about abortions, contact experiences with people seeking an abortion, fear of stigmatization (if participating in abortion care in the future), empathy, subjective norms, perceived behavioral control, and willingness to participate in abortion care. Abortion attitudes were assessed via the AAS.

The cross-sectional online study including the secondary analysis at hand was preregistered on AsPredicted (registration number 142964, https://aspredicted.org/b5b2q.pdf).

### Measure

The 14-item AAS was first developed to be used by health educators helping students to explore their values concerning abortion. The author validated the scale by assessing interrater agreement, and reliability estimates. Factor analysis provided a unidimensional structure (33). When applying the scale to high school and college students, active “Right to Life” members, and abortion service associates, the AAS showed a high reliability (α = .92) and high construct validity (33). Furthermore, the instrument could clearly differentiate between “Right to Life” members and abortion service associates (33).

Items of the AAS are rated on a Likert-scale from 0 (“strongly disagree”) to 5 (“strongly agree”) (33). For the original English items of the 14-item scale, see Table 1. For calculating a sum score, the scales of items 1, 3, 4, 7, 9, 12, and 14 have to be reversed. Higher AAS scores indicate a stronger pro abortion attitude. According to Sloan (1983), participants can be grouped to one of five categories according to their sum score: strong contra abortion (original wording: strong pro life, sum score 0-15), moderate contra abortion (original wording: moderate pro life, sum score 16-26), unsure (sum score 27-43), moderate pro abortion (sum score 44-55), strong pro abortion (sum score 56-70).

**Table 1:** Comparison of the original AAS scale and the German AAS scale provided in a translated version.

|  | Original scale | German AAS [translated]* |
| --- | --- | --- |
| Introduc<br>tion | This is not a test. There are no wrong or right answers to any of the statements, so just answer as honestly as you can. The statements ask you to tell how you feel about legal abortion (the voluntary removal of a human fetus from the mother during the first three months of pregnancy by a qualified medical person). Tell how you feel about each statement by circling one of the choices beside each sentence. Here is a practice statement:<br>Abortions should be legalized.<br>strongly agree - agree - slightly agree - slightly disagree - disagree - strongly disagree<br>Please respond to each statement and circle only one response. No one else will see your responses without permission. | This is not a test. There are no wrong or right answers to any of the statements, so just answer as honestly as you can. The statements are about your views on **abortion during the first three months of pregnancy by a qualified health professional. Please indicate how much you agree or disagree with a statement by checking an answer option.** Here is an example:<br>Abortions should be legalized.<br>strongly agree - agree - slightly agree - slightly disagree - disagree - strongly disagree<br>Please respond to each statement and **select only one response** at a time. No one else will see your responses without permission. |
| Item 1 | The supreme court should strike down legal abortions in the United States. | **In Germany, abortions should be banned under all circumstances.** |
| Item 2 | Abortion is a good way of solving an unwanted pregnancy. | Abortion is a good way of solving an unwanted pregnancy. |
| Item 3 | A mother should feel obligated to bear a child she has conceived. | **A person** should feel obligated to bear a child **that was conceived.** |
| Item 4 | Abortion is wrong no matter what the circumstances are. | **Abortions** are wrong no matter what the circumstances are. |
| Item 5 | A fetus is not a person until it can live outside its mother's body. | A fetus is not a person until it can live outside **the pregnant person's** body. |
| Item 6 | The decision to have an abortion should be the pregnant mother's. | The decision **for an abortion** should be the **pregnant person's.** |
| Item 7 | Every conceived child has the right to be born. | Every conceived child has the right to be born. |
| Item 8 | A pregnant female not wanting to have a child should be encouraged to have an abortion. | A **pregnant person** not wanting to have a child should be encouraged to have an abortion. |
| Item 9 | Abortion should be considered killing a person. | Abortion should be considered killing a person |
| Item 10 | People should not look down on those who choose to have abortions. | People should not look down on those who choose to have **an abortion.** |
| Item 11 | Abortion should be an available alternative for unmarried, pregnant teenagers. | Abortion should be an **easily accessible alternative for pregnant minors.** |
| Item 12 | Persons should not have the power over the life or death of a fetus. | **No one** should have the power over the life or death of a fetus. |
| Item 13 | Unwanted children should not be brought into the world. | Unwanted children should not be brought into the world. |
| Item 14 | A fetus should be considered a person at the moment of conception. | A fetus should be considered a person at the moment of conception. |

Note: Changes of the German scale compared to the English scale are marked in bold letters. Translation of the German AAS scale into English was done by the first author alone without applying a team translation approach and only for the purpose of readability within this manuscript. The German AAS translated into English is not a systematic back translation, not validated and should therefore not be used without further testing.

### Translation

For translating and adapting the English AAS into German, we utilized the team translation protocol TRAPD (Translation, Review, Adjudication, Pretesting, and Documentation, (38–40). Initially, two authors (AL, MB) who are fluent in both German and English independently translated the English AAS into German. Furthermore, they suggested adaptations of items, if necessary due to cultural or legal differences of the (political) context. Next, a third team member (MR, see List of Abbreviations) reviewed the translations, selecting one version or proposing a third version. In a final discussion, AL, MB, and MR reached an agreement on the translated and adapted German version.

### Assessment of comprehensibility

Following the COSMIN criteria (Consensus-based Standards for the Selection of Health Measurement Instruments) (41)], we assessed comprehensibility by evaluating whether the AAS items are understood by medical students as intended. Thereby, we conducted three rounds of cognitive interviews with a convenience sample of n=10 medical students currently studying human medicine at a medical school in Germany. The interviews, led by MB, were conducted online via Zoom and audio-recorded. Participants’ demographic characteristics were assessed and descriptive statistics were calculated using SPSS (IBM SPSS Statistics, Version 29.0.1.0). An interview guide was created based on the recommendations of Willis et al. (42), incorporating verbal probing techniques and paraphrasing. Following each round of cognitive interviews, AL and MB analyzed and discussed the feedback and suggestions from the participants. We revised the German AAS items accordingly and tested the revised versions in the next round.

We furthermore send the final survey of the cross-sectional study (including the AAS), to our cooperation partners, who supported the study team during their research on abortion care. We received feedback from n=2 abortion counselors and n=3 gynecologists providing abortion care. According to their feedback, further modification of the AAS were not necessary.

### Data collection

Data for the cross-sectional study were collected using Unipark (EFS Survey, Tivian XI GmbH) between September 8^th^ and October 4^th^ 2023. We included medical students, who were at least 18 years old enrolled at a medical school in Germany at the time of participation. Medical student councils of all medical faculties in Germany were contacted via email and asked to resend the study invitation and a link to the survey to their students or via online postings on the faculty bulletin boards. Additionally, local groups of Medical Students for Choice (MSFC), an international initiative to support safe abortions, distributed the survey link via internal faculty email lists and Instagram channels. Participation was voluntary and without financial incentives. Participants were informed about the study’s purpose and background, as well as the procedure for data storage and analysis. They provided informed consent by ticking the respective box after reading the study information. The AAS was the first instrument participants had to fill out within the survey, followed by several other items and instruments (24). Demographic characteristics were assessed at the end of the survey.

### Psychometric data analyses

We excluded data sets, if 1) they were completely empty, 2) had more than 30% missing AAS items (43), 3) participants did not fulfil the inclusion criteria, and / or 4) answered one of the two control questions wrong. Those control questions were used to detect careless responses (44,45). We furthermore screened data sets for suspicious response behaviour (e.g., participant always choose the same response option) and excluded suspicious data sets after discussions within the study team.

For the included participants, we calculated descriptive statistics of their demographic characteristics. For analysis of acceptance, we analyzed the completion rate (percentage of missing values) per item.

In the next step, we conducted item analyses. This involved examining response distribution, item difficulty (calculated by standardizing the mean to range from 0 to 1, recommended range 0.2 – 0.8) to assess floor and ceiling effects (item difficulties below 0.2 indicate a floor effect; item difficulties above 0.8 indicate a ceiling effect). Additionally, we calculated item means and standard deviations, corrected item-total correlations (recommended range >0.3), inter-item correlations (recommended range >0.3), and sum scores (46–48). For analysis of corrected item-total correlations and inter-item correlations, missing data were replaced by multiple imputation (49).

To assess structural validity, we conducted a confirmatory factor analysis (CFA). First, we examined the suitability of factor analysis, by analyzing the Kaiser-Meyer-Olkin (KMO) measure of sampling adequacy and Bartlett’s test of sphericity (50,51). KMO value should be higher than .05 and Bartlett’s test value should be less than .05 to fulfil the criteria for calculating a factor analysis (50,51). The author of the original AAS postulated a one-factor structure of the AAS (33). However, the original publication lacks information on factor loadings or fit indices for this assumption. Furthermore, other studies using the AAS did not analyze the factorial structure of the instrument but relied on the assumption of a one-factor-structure (34,35). Thus, we initially hypothesized a one-factor structure for the German AAS and modified the model step-by-step based on the results of the factor analysis, item analysis and content of specific items. We conducted a CFA with a unidimensional model (model 1) (52–54) using a robust version of maximum likelihood estimator (MLR) and full information maximum likelihood to address missing values. There are various criteria for interpreting factor loadings and determining cut-offs for appropriate factor models (55,56). For this study, we chose to use the established, less conservative criterion of .40 as the cut-off for acceptable factor loadings in samples larger than n=200 (55). To evaluate model fit of each model, a range of global goodness-of-fit indices were calculated: discrepancy chi-squared statistic (Chi^2^), Comparative Fit Index (CFI), Tucker-Lewis Index (TLI), Root Mean Square Error of Approximation (RMSEA), and Standardized Root Mean Squared Residual (SRMR). For comparison of models we used Akaike information criterion (AIC) and Bayesian Information Criterion (BIC). We used established criteria to interpret the fit of the estimated models (57,58). Afterwards, we assessed reliability by McDonald’s omega (59).

Analysis of demographic data, completion rate, and item analysis were done using SPSS (IBM SPSS Statistics, Version 29.0.1.0). We used R Version 4.3.2 (R Core Team, Vienna, Austria) for CFA (specifically, lavaan package (60)) and reliability analyses (specifically, semtools (61)).

## RESULTS

### Translation and adaptation

Both translators (AL and MB) and the reviewer (MR) had similar translations of the ASS. S1

Appendix (Table A) provides an overview on the original AAS, results after translation and adaptation as well as the final German instrument. Within the first round of team discussion, we reached consensus for translation and adaptation of all items, the response scale, and the introduction. In the introduction, we decided to remove the term “legal” from “legal abortions” because abortions are illegal under the German criminal code (but will not be punished under certain conditions, §218 StGB (62)). Item 1 had to be adapted to the German policy and national legal restrictions. Here, the term “legal abortions” might also be misleading in the German context, so we decided for the term “under all circumstances” (dt. “unter allen Umständen”). For item 5, we added an alternative version avoiding the negation to increase comprehensibility of the item. For adaptation of item 6, we added an alternative version, which slightly differ in its word order. We discussed item 9 regarding the meaning of the original term “killing a person”. In German, the two phrases “killing” and “murder” (“Tötung/Totschlag” and “Mord”) have different definitions according to the German criminal code (§211 and §212 StGB (63)). Those are comparable to the American legal system but there, their definitions are not laid down in a uniform set of rules but in respective criminal statutes of each state. Thus, we decided to discuss both versions “killing” and “murder” with our participants. We also tested two versions of item 10 containing the singular and plural form of the term “abortion”. Item 11 had also be adapted culturally since the public view on the relevance of marital status of “unmarried pregnant teenager” has changed within the past 40 years and might not be comparable between Germany and the United States. Additionally, abortion actually is an option for pregnant teenager by German law. We decided to test two versions, which differ slightly in wording and the use of the term “unmarried”. We adapted item 12 by changing the term “persons” (dt. “Personen”) to “nobody” (dt. “niemand”), to avoid negation and improve readability. Finally, to make the instrument more inclusive, we changed the term “mother” or “female” to “(pregnant) person” in item 3, item 5, item 6, and item 8. Pregnant person should also address trans men and other individuals, who would not call themselves a mother or female person but are able to become pregnant. Finally, we decided to change two results categories from “moderate / strong pro life” to “moderate / strong contra abortion” to avoid reproducing nomenclature used by anti-abortion activists.

### Assessment of comprehensibility

We conducted cognitive interviews with n=10 medical students. For demographic data of participants, see Table B in S1 Appendix.

The first round of cognitive interviews (n=3) revealed that the adapted introduction, the response scale and item 2, item 4, item 7, item 12, and item 14 were well understood. However, those items were tested again in round 2 (n=2) and 3 (n=5) of cognitive interviews but were still understood well by all participants and accordingly had not to be adapted. We decided to test another version for item 1 in round 2 and 3 (using the term “basically” (dt. “grundsätzlich”) instead of “under all circumstances” (dt. “unter allen Umständen”) Participants discussed that “basically” is the “weaker” term compared to “under all circumstances” by not including all possible circumstances. To be as close as possible to the original item, the study team decided to use “under all circumstances”. After interview round 1, another version of item 3 using a passive term was tested (“she has conceived” / dt. “das sie gezeugt hat” vs. “who was conceived” / dt. “das gezeugt wurde”). Participants of the following two rounds preferred the second version. Almost all participants preferred the second version of the adapted item 5 and 6 since it was better comprehensible by avoiding negation and an interlaced sentence. After the first round of cognitive interviews and based on suggestions by participants, we developed a new version for item 8 including the term “should be supported in having an abortion” (dt. “sollte bei einem Schwangerschaftsabbruch unterstützt werden”) compared to “should be encouraged to have an abortion” (dt. “sollte zu einem Schwangerschaftsabbruch ermutigt werden“). Participants of round 2 and 3 discussed that this version has a more “active” and “directive” meaning. Thus, the study team decided for the first translated and adapted version to stick to the original item. Version 1 of item 9 was preferred by almost all participants because the term “killing” (dt. “Tötung/Totschlag”) was described as less harmful compared to “murder” (dt. “Mord”) and might therefore be less stigmatizing. Since the translation of the term “killing” is also closer to the original item, the study team decided to use the first version of the adapted item 9 with slight modifications in wording. For item 10, participants preferred the adapted item version 2, which uses the singular form of “abortion”. Thus, the study team decided to use this version, even though it is less congruent with the original item. Based on discussions about item 11 within the first round of cognitive interviews, the study team decided to again adapt the item and test a new item version, which is a mixture of the previously tested versions 1 and 2. This third version was well understood by all participants of interview round 2 and 3 and thus used for the final instrument. After the first round of cognitive interviews and suggestions by participants, we developed a new version of item 13, which included the term “should not be given birth to” (dt. “sollten nicht auf die Welt gebracht werden”) instead of “should not be brought into the world” (dt. „sollten nicht in die Welt gesetzt werden”), since the new version is less offensive. This new version was well understood by participants of round 2 and 3 and thus used for the final instrument. The final German AAS instrument can be found in S1 Appendix (Table A), a translated version is displayed in Table 1.

### Psychometric evaluation

#### Data cleaning

Before analysis, 46 data sets were excluded because they were empty (n=35), the inclusion criteria were not fulfilled (n=6), the second control question was incorrectly answered (n=3), and identical and suspicious responses for most of the scales (n=2). Data of 305 medical students were available for further analysis.

#### Sample characteristics

Table 2 gives an overview of participants’ demographic characteristics. Mean age of the 305 included medical students was 23.7 years (SD 3.81) and on average they were in the 6^th^ semester (SD 3.08). Most were female (63.3%), living in a permanent unmarried partnership (37.7%), currently live in the state North Rhine-Westphalia (44.9%) and their preferred specialization for the future was gynecology and obstetrics (11.8%). Most have no religious beliefs (38.4%) and never visit a religious site (31.1%). 11 participants (3.6%) already had an abortion themselves. Further details on participants’ demographic characteristics (own country of birth, parents country of birth, state of origin, hometown size, mother tongue, pregnancies, children, and confession) can be found in S2 Appendix (Table A).

**Table 2:** Demographic characteristics of participants (n=305 medical students).

|  |  |  |  |
| --- | --- | --- | --- |
| Age | **Mean** | 23.7 |  |
|  | **SD** | 3.81 |  |
|  | **n** | 234 |  |
| Semester | **Mean** | 6.67 |  |
|  | **SD** | 3.08 |  |
|  | **n** | 234 |  |
|  |  | **n** | **%** |
| Gender | Female | 193 | 12.5 |
|  | Male | 38 | 63.3 |
|  | Divers / non-binary | 1 | 0.3 |
|  | Preferred not to answer this question | 2 | 0.7 |
|  | Missings | 71 | 23.3 |
| Relationship status | Single | 105 | 34.4 |
|  | Permanent partnership (unmarried) | 115 | 37.7 |
|  | Married | 12 | 3.9 |
|  | Divorced | 2 | 0.7 |
|  | Missings | 71 | 23.3 |
| Current state of living | Baden-Wuerttemberg | 16 | 5.2 |
|  | Bavaria | 12 | 3.9 |
|  | Berlin | 2 | 0.7 |
|  | Hamburg | 6 | 2.0 |
|  | Hesse | 1 | 0.3 |
|  | Lower-Saxony | 16 | 5.2 |
|  | North Rhine-Westphalia | 137 | 44.9 |
|  | Saxony | 38 | 12.5 |
|  | Schleswig-Holstein | 5 | 1.6 |
|  | Thuringa | 1 | 0.3 |
|  | Missings | 71 | 23.3 |
| Had an abortion themselves | 0 | 219 | 71.8 |
|  | 1 | 11 | 3.6 |
|  | Missings | 75 | 24.6 |
| Future preferred specialization | General medicine | 28 | 9.2 |
|  | Anesthesiology | 28 | 9.2 |
|  | Anatomy | 1 | 0.3 |
|  | Ophthalmology | 5 | 1.6 |
|  | Dermatology | 3 | 1.0 |
|  | Gynecology and obstetrics | 36 | 11.8 |
|  | Otolaryngology | 2 | 0.7 |
|  | Hematology | 1 | 0.3 |
|  | Internal medicine | 23 | 7.5 |
|  | Cardiology | 7 | 2.3 |
|  | Pediatrics and adolescent medicine | 34 | 11.1 |
|  | Child and adolescent psychiatry and psychotherapy | 3 | 1.0 |
|  | Oral and maxillofacial surgery | 5 | 1.6 |
|  | Neurosurgery | 12 | 3.9 |
|  | Neurology | 10 | 3.3 |
|  | Public health care | 1 | 0.3 |
|  | Oncology | 6 | 2.0 |
|  | Pathology | 2 | 0.7 |
|  | Pharmacology | 1 | 0.3 |
|  | Psychiatry and psychotherapy | 6 | 2.0 |
|  | Psychosomatic medicine and psychotherapy | 1 | 0.3 |
|  | Radiologie | 8 | 2.6 |
|  | Forensic medicine | 6 | 2.0 |
|  | Urology | 3 | 1.0 |
|  | Missing | 73 | 23.9 |
| Influence of faith | I have no religious beliefs | 117 | 38.4 |
|  | Not at all | 36 | 11.8 |
|  | Low | 43 | 14.1 |
|  | Somewhat | 19 | 6.2 |
|  | Rather strong | 15 | 4.9 |
|  | Very strong | 4 | 1.3 |
|  | Missings | 71 | 23.3 |
| Visit of a religious sites | Never | 95 | 31.1 |
|  | Once a year | 61 | 20.0 |
|  | Several times a year | 59 | 19.3 |
|  | Several times a month | 7 | 2.3 |
|  | Once a week or more often | 11 | 3.6 |
|  | Missings | 72 | 23.6 |

Note: SD = standard deviation

Most participants could be grouped to the category “Strong pro abortion” (47.87%) or “Moderate pro abortion” (36.39%), 12.3% were grouped to the category “Unsure” and a minority showed “Moderate contra abortion” (2.9%) or “Strong contra abortion” (0.6%) attitudes (see Table 3).

**Table 3:**
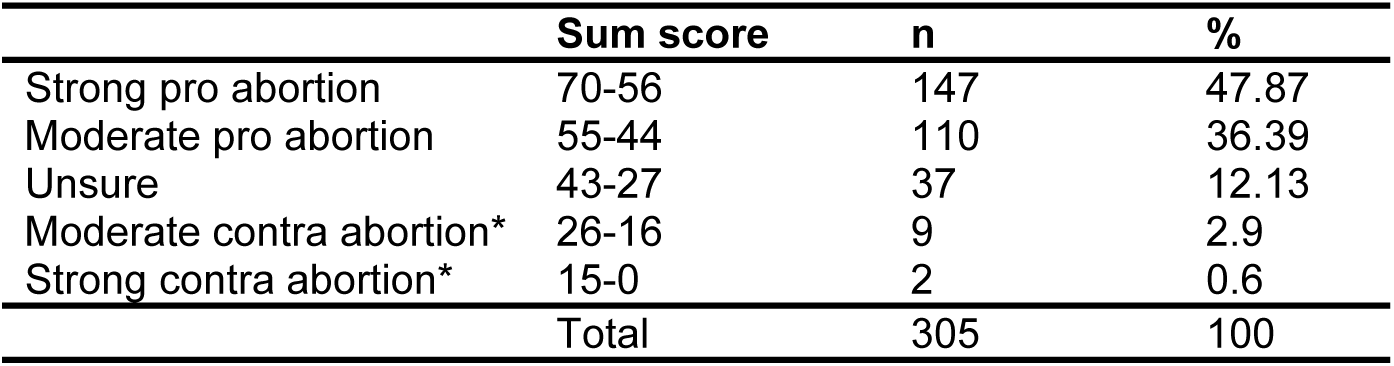
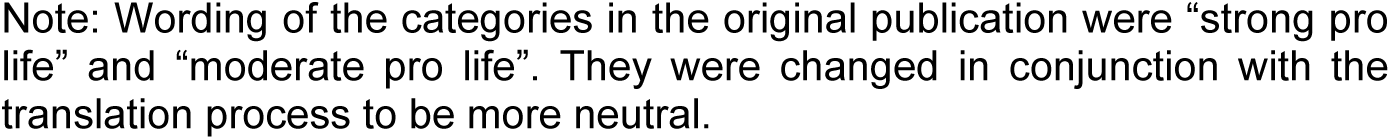
Sum scores and categories for the German AAS.

|  | Sum score | n | % |
| --- | --- | --- | --- |
| Strong pro abortion | 70-56 | 147 | 47.87 |
| Moderate pro abortion | 55-44 | 110 | 36.39 |
| Unsure | 43-27 | 37 | 12.13 |
| Moderate contra abortion* | 26-16 | 9 | 2.9 |
| Strong contra abortion* | 15-0 | 2 | 0.6 |
| Total |  | 305 | 100 |

Note: Wording of the categories in the original publication were “strong pro life” and “moderate pro life”. They were changed in conjunction with the translation process to be more neutral.

##### Acceptance

All items had high acceptance. The highest rate of missing values was 4 missing values (1.32%) for Item 12 (see Table 4). Missing values could be observed for 9 cases with one missing value per case. Accordingly, 97.05% of the respondents filled out the AAS completely.

**Table 4:** Means, standard deviation, skewness, item difficulty, acceptance and item discrimination of the German AAS (n=305 medical students).

|  | Strongly<br>Disagree<br>N (%) | Disagree<br>N (%) | Slightly<br>Disagree<br>N (%) | Slightly<br>Agree<br>N (%) | Agree<br>N (%) | Strongly<br>Agree<br>N (%) | Mean (SD) | Item<br>difficulty | Acceptance<br>(Completion<br>rate in %) | Corrected<br>item-total<br>correlation<br>Model 1 | Corrected<br>item-total<br>correlation<br>Model 2 |
| --- | --- | --- | --- | --- | --- | --- | --- | --- | --- | --- | --- |
| Item 1 | 269 (88.2%) | 19 (6.2%) | 7 (2.3%) | 4 (1.3%) | 4 (1.3%) | 1 (0.3%) | 4.78 (0.72%) | 0.96 | 99.67 | 0.552 | 0.558 |
| Item 2 | 7 (2.3%) | 6 (2.0%) | 14 (4.6%) | 40 (13.1%) | 75 (24.6%) | 163 (53.4%) | 4.16 (1.17%) | 0.83 | 100.0 | 0.719 | 0.721 |
| Item 3 | 188 (61.6%) | 47 (15.4%) | 33 (10.8%) | 21 (6.9%) | 10 (3.3%) | 6 (2.0%) | 4.19 (1.26%) | 0.84 | 100.0 | 0.697 | 0.696 |
| Item 4 | 251 (82.3%) | 35 (11.5%) | 10 (3.3%) | 4 (1.3%) | 3 (1.0%) | 2 (9.7%) | 4.71 (0.78%) | 0.94 | 100.0 | 0.523 | 0.525 |
| Item 5 | 31 (10.2%) | 58 (19.0%) | 77 (25.2%) | 59 (19.3%) | 56 (18.4%) | 24 (7.9%) | 2.40 (1.44%) | 0.48 | 100.0 | 0.594 | 0.592 |
| Item 6 | 1 (0.3%) | 3 (1.0%) | 6 (2.0%) | 33 (10.8%) | 78 (25.6%) | 184 (60.3%) | 4.41 (0.87%) | 0.88 | 100.0 | 0.644 | 0.637 |
| Item 7 | 79 (25.9%) | 80 (26.2%) | 76 (24.9%) | 46 (15.1%) | 10 (3.3%) | 13 (4.3%) | 3.44 (1.34%) | 0.68 | 99.67 | 0.750 | 0.754 |
| Item 8 | 41 (13.4%) | 54 (17.7%) | 112 (36.7%) | 67 (22.0%) | 23 (7.5%) | 7 (2.3%) | 1.99 (1.21%) | 0.39 | 99.67 | 0.462 | 0.460 |
| Item 9 | 207 (67.9%) | 48 (15.7%) | 22 (7.2%) | 17 (5.6%) | 5 (1.6%) | 6 (2.0%) | 4.37 (1.14%) | 0.87 | 100.0 | 0.770 | 0.766 |
| Item 10 | 10 (3.3%) | 0 (0.0%) | 3 (1.0%) | 14 (4.6%) | 38 (12.5%) | 240 (78.7%) | 4.59 (1.03%) | 0.92 | 100.0 | 0.295 | - |
| Item 11 | 11 (3.6%) | 22 (7.2%) | 44 (14.4%) | 68 (22.3%) | 65 (21.3%) | 95 (31.1%) | 3.44 (1.43%) | 0.69 | 100.0 | 0.640 | 0.642 |
| Item 12 | 119 (39.0%) | 97 (31.8%) | 63 (20.7%) | 15 (4.9%) | 3 (1.0%) | 4 (1.3%) | 4.00 (1.05%) | 0.80 | 98.68 | 0.706 | 0.711 |
| Item 13 | 34 (11.1%) | 58 (19.0%) | 70 (23.0%) | 85 (27.9%) | 36 (11.8%) | 20 (6.6%) | 2.30 (1.38%) | 0.46 | 99.33 | 0.558 | 0.562 |
| Item 14 | 129 (42.3%) | 69 (22.6%) | 65 (21.3%) | 28 (9.2%) | 6 (2.0%) | 8 (2.6%) | 3.86 (1.26%) | 0.77 | 100.0 | 0.678 | 0.672 |

Note: Values for Items 1, 3, 4, 7, 9, 12 and 14 are reversed to calculate means and item difficulties. Corrected item-total correlations were calculated for model 1 (with all items and no correlations between items) and model 2 (without item 10 but no correlations between items).

##### Analysis of AAS items

Table 4 shows response distribution, means and standard deviations, item difficulties, acceptance (completion rates), and corrected item-total correlations of the 14 items. Means ranged between 1.99 (item 8) and 4.78 (item 1) on a scale from 0 to 5. Accordingly, item difficulties ranged from 0.39 (item 8) to 0.96 (item 1) with 7 items exceeding 0.8 and thus demonstrating ceiling effects. Corrected item-total correlations ranged from 0.295 (item 10) to 0.770 (item 9) for model 1 and 0.469 (item 8) to 0.766 (item 9) for model 2, and inter-item correlations from .129 (item 1 / item 10) to .681 (item 7 / item 12, see S2 Appendix, Table B).

##### Factor analysis

Assumptions for factor analysis were met (41,50,51). KMO measure was .932 and Bartlett’s test of sphericity yielded χ^2^ = 2016.04, p < .001 (50,51). For model 1, we assumed a one-factorial structure without correlations between items based on the evaluation of the original instrument (33). CFA for this model showed standardized factor loadings between .305 (item 10) and .828 (item 9) with one item showing a factor loading below .40 (item 10; see Table 2). This indicated that item 10 did not fit to the predefined factor (55,56,64). Values of RMSEA and TLI did not meet cut-offs (65,66) (see Table 3). We analysed item 10 on the content level. In comparison to the other items, item 10 does not refer to people who have abortions, but to other people who make negative judgments about people having abortions. This item seems to measure the attitudes towards abortion rather implicitly. Discussing those results together with results of the item analysis for item 10, we decided to calculate another alternative model without item 10 (model 2). Model 2 had factor loadings between 0.437 (item 8) and 0.827 (item 9) (see Table 2). Model fits for model 2 were comparable to model 1 with values of RMSEA and TLI below cut-off. However, AIC and BIC showed better model fit for model 2 (see Table 3). We furthermore calculated modification indices and found that the χ^2^ test statistic for freeing the covariance between Item 8 and Item 13 was 45.519 (degree of freedoms = 1). Furthermore, we discussed in the study team a strong similarity of the items content since both items contain the same statement, only described in other words. Accordingly, we calculated a model 3 with a correlation between item 8 and item 13 and found factor loadings between 0.418 and 0.819 (see Table 2). Model fits for model 3 were better compared to model 1 and 2.

**Table 2:**
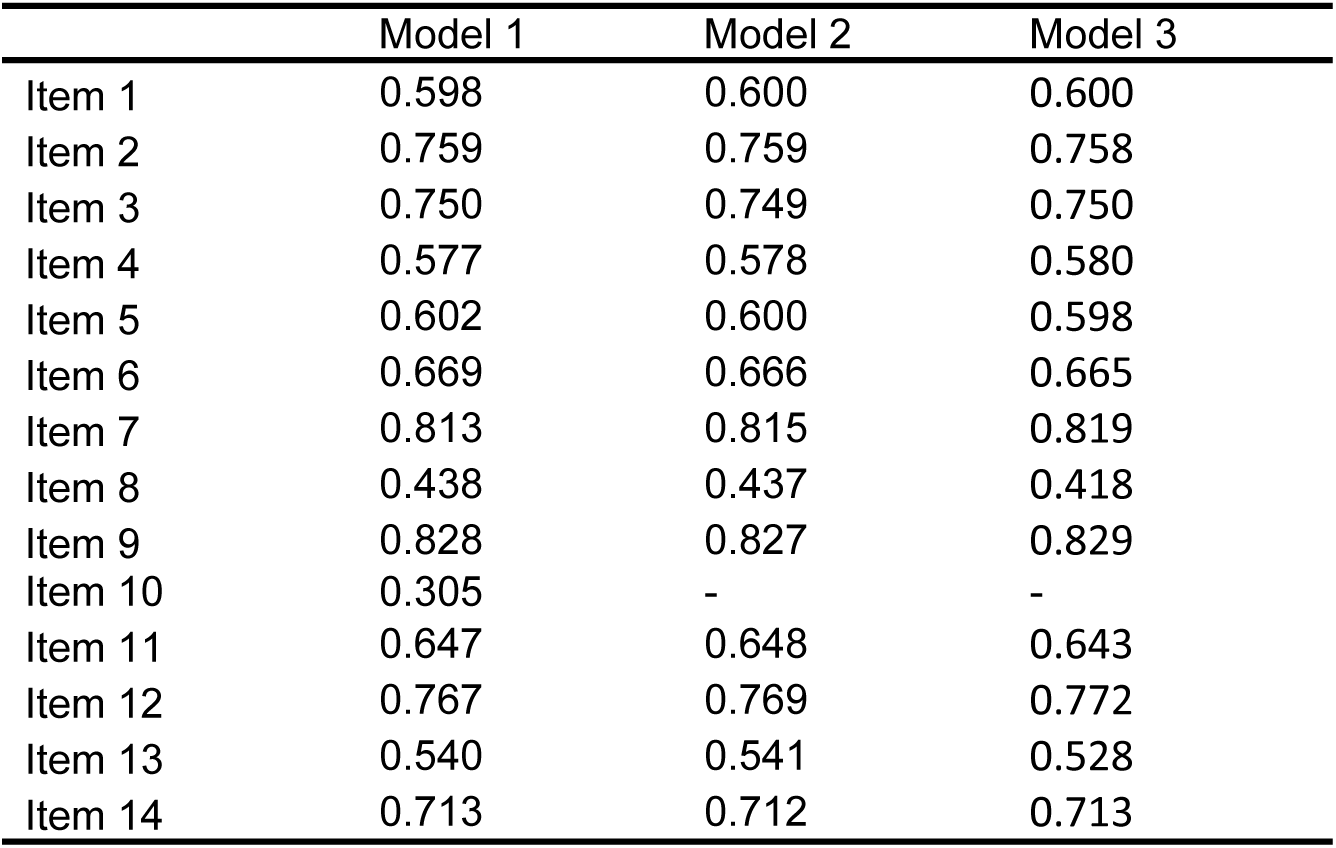
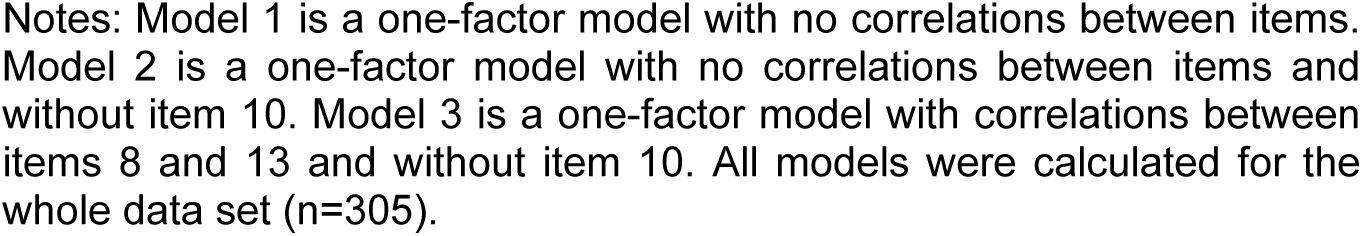
Factor loadings of two calculated models for the factor analysis of the German AAS (n=305 medical students).

|  | Model 1 | Model 2 | Model 3 |
| --- | --- | --- | --- |
| Item 1 | 0.598 | 0.600 | 0.600 |
| Item 2 | 0.759 | 0.759 | 0.758 |
| Item 3 | 0.750 | 0.749 | 0.750 |
| Item 4 | 0.577 | 0.578 | 0.580 |
| Item 5 | 0.602 | 0.600 | 0.598 |
| Item 6 | 0.669 | 0.666 | 0.665 |
| Item 7 | 0.813 | 0.815 | 0.819 |
| Item 8 | 0.438 | 0.437 | 0.418 |
| Item 9 | 0.828 | 0.827 | 0.829 |
| Item 10 | 0.305 | - | - |
| Item 11 | 0.647 | 0.648 | 0.643 |
| Item 12 | 0.767 | 0.769 | 0.772 |
| Item 13 | 0.540 | 0.541 | 0.528 |
| Item 14 | 0.713 | 0.712 | 0.713 |

Notes: Model 1 is a one-factor model with no correlations between items. Model 2 is a one-factor model with no correlations between items and without item 10. Model 3 is a one-factor model with correlations between items 8 and 13 and without item 10. All models were calculated for the whole data set (n=305).

**Table 3:** Fit indices of two calculated models for factor analysis of the German AAS (n=305 medical students).

|  | Chi<sup>2</sup> <sup>1</sup> | df <sup>2</sup> | p value | RMSEA <sup>3</sup> | SRMR <sup>4</sup> | TLI <sup>5</sup> | CFI <sup>6</sup> | AIC <sup>7</sup> | BIC <sup>8</sup> | Omega |
| --- | --- | --- | --- | --- | --- | --- | --- | --- | --- | --- |
| Recommen-<br>dation |  |  | >0.05 | ≤0.06 | <0.08 | ≥0.90 | ≥0.90 | Model with the<br>smallest value |  |  |
| Model 1 | 252.1 | 77 | <0.001 | 0.086 | 0.051 | 0.895 | 0.911 | 11342 | 11498 | 0.910 |
| Model 2 | 235.9 | 65 | <0.001 | 0.093 | 0.053 | 0.894 | 0.912 | 10483 | 10628 | 0.913 |
| Model 3 | 187.4 | 64 | <0.001 | 0.079 | 0.045 | 0.923 | 0.937 | 10437 | 10585 | 0.904 |

Notes: Model 1 is a one-factor model with no correlations between items. Model 2 is a one-factor model with no correlations between items and without item 10. Model 3 is a one-factor model with correlations between items 8 and 13 and without item 10. All models were calculated for the whole data set (n=305). <sup>1</sup> Discrepancy chi-squared statistic, <sup>2</sup> Degrees of freedom, <sup>3</sup> Root mean square error of approximation, <sup>4</sup> Standardized Root Mean Squared Residual, <sup>5</sup> Tucker-Lewis Index, <sup>6</sup> Comparative Fit Index, <sup>7</sup> Akaike Information Criterion, <sup>8</sup> Bayesian Information Criterion.

## DISCUSSION

The original AAS is a brief instrument to assess abortion attitudes of students and might also be applicable for the community. It was developed and evaluated in 1983 in the Unites States (33). Our study aimed to translate the English AAS into German, adapt it to the specific societal and legal context of Germany and evaluate its psychometric properties.

### Translation, adaptation and psychometric evaluation

The translation team quickly reached consensus for the translation of the AAS. No adaptations of the translated versions were necessary for items 2, 4, 7, 13, and 14. After translation and, if necessary, adaption by the study team, the introduction as well as items 1, 2, 4, 7, 8, 12, and 14 were well understood by all participants of cognitive interviews. For items 5, 6, and 10, participant decided in favour of one of two item versions developed by the study team. We reworded items 3, 9, 11, and 13 based on feedback of participants of cognitive interviews and new item versions were included in the final instrument. The last round of cognitive interviews demonstrated that comprehensibility of all final items was given.

All items of the German AAS showed high acceptability. The majority of participants showed to have high AAS sum scores and were categorized having a strong pro or moderate pro abortion attitude (47.87% and 36.39% respectively). Only 3.5% were categorized having moderate or strong contra abortion attitudes. In our data ceiling effects of most of the items became evident, reflecting the highly positive abortion attitude of our specific sample. Hollis and Morris found that, when asked about their abortion attitudes, most of their participants clustered at the low end or the high end even when applying a 7-point Likert scale to increase variance of responses (67). However, response distribution in our data does not reflect the results of Swartz et al. (2020), who assessed the AAS in 504 nurses in California (United States) and found rather negative attitudes towards abortion (68). They did not calculate sums scores but found that, for example, 52.4% (n=264) strongly agreed to item 4 (“Abortion is wrong no matter what the circumstances are.”) compared to 0.97% (n=2) in our sample. In our sample, 82.3% (n=251) strongly disagreed with this item. However, item analysis of data of Swartz et al. disclose some contradictions (68). For example, in their sample, 53.8% (n=271) (strongly/somewhat) agreed that every conceived child has the right to be born (item 7) but 33.7% (n=397) (strongly/somewhat) agreed that a pregnant women who does not want a child should be encouraged to have an abortion (item 8). Alspaugh et al. (2022), who assessed abortion attitudes in women’s health and neonatal nurses in the United States, found a more diverse picture (35). In a sample of 1.820 nurses, 16% had strong pro abortion attitudes, 32% had moderate pro abortion attitudes, 29% were unsure, 11% had moderate contra abortion attitudes and 13% had strong contra abortion attitudes. In our sample of German medical students, most preferred to work in the field of gynecology and obstetrics later. It can be assumed that based on the current political and social discussions about abortion regulations in Germany, our sample was rather motivated to participate because of their pro abortion attitude, has therefore a self-selection bias and might not be representative for other groups. However, results of all three studies disclose that the AAS has the potential to differentiate between pro and contra abortion attitudes and is sensitive to the specific demography and background of different samples within a community.

Item characteristics for the German AAS was satisfying except for item 10 (original: “People should not look down on those who choose to have abortions”), which showed corrected item-total correlations below cut-off and additionally sub-optimal values for response distribution (demonstrating a ceiling effect), and inter-item correlation. This indicates that item 10 does not measure the underlying concept (50,51,69). When calculating model 1, the a priory hypothesized one-factorial structure for the German AAS was confirmed but item 10 was the only item with a factor loading below the cut-off of 0.3. Also, model fits were not fully satisfying. After removing item 10 based on discussions on item analysis and content of the item (model 2), we also decided to allow correlations between item 8 and item 13 based on modification indices and discussions about the items content (model 3). The resulting model 3 was the most satisfying with acceptable model fits and factor loadings (55,56,64). For future use of the German AAS we recommend to remove item 10 from the instrument and psychometrically evaluate this 13-item German AAS.

### Broader implications of the AAS

The World Health Organization considers abortion as a key part of comprehensive sexual and reproductive health care (70). However, physicians’ attitudes toward it have been largely overlooked. The successful translation and adaptation of the AAS into German highlights its potential as a culturally sensitive tool for evaluating abortion attitudes in different sociocultural contexts. This adaptability is crucial for understanding how societal norms, laws, and individual beliefs intersect to shape abortion attitudes and abortion stigma in diverse populations. In countries like Germany, where abortion care is politically and ethically sensitive, the AAS could assess the attitudes of medical students and healthcare providers, informing educational interventions aimed at promoting unbiased, person-centered care. Understanding attitudes early in medical training could also help identify potential barriers to providing comprehensive abortion care and address them through targeted curricula (15,24,71). A tool like the AAS enables researchers, clinicians and policymakers to explore how community attitudes affect women’s mental health, decision-making, and experiences of stigma, particularly in contexts where restrictive laws and cultural norms contribute to feelings of shame, guilt, or fear among women seeking abortions. Additionally, the AAS could be employed in longitudinal studies to track changes in abortion attitudes over time, especially in countries undergoing significant legal or cultural shifts regarding reproductive rights. This would allow to measure the impact of these changes on public opinion.

### Strengths and Limitations

A major strength of this study is that we provided the first measure to assess abortion attitudes in German language to be used in diverse settings and groups. We applied an established state-of-the-art translation procedure and used a qualitative approach to explore comprehensibility. Furthermore, we applied the German version of the AAS in a sample, which was large enough to robustly perform psychometric analysis. While the German AAS has shown promise, its limitations — such as the need to remove item 10 — highlight the importance of ongoing validation and refinement. In our study, a self-selection bias of participants who are interested in the topic cannot be ruled out. Future studies should explore its application in more diverse and representative samples, including non-medical populations, healthcare providers, and patients, to ensure its broad utility and relevance. Furthermore, we had a high number of missing data within the demographic data of our participants, since those were assessed at the end of the survey. This limits the precise description of our sample. Since this was a secondary analysis of cross-sectional data, several psychometric parameters were not analysable for our data. It was for example not possible to assess convergent or divergent validity. In further validation studies, convergent validity of the AAS could be tested by also applying the recently published 7-item Community Attitudes Abortion Scale (CAAS), which was developed from a survey of 1.533 women without a history of abortion seeking family planning services in the United States. It measures attitudes toward women who have abortions, anticipated behaviour towards a friend who has had an abortion, and attitudes towards abortion regulation (72).

## CONCLUSION

It is essential to measure abortion attitudes of medical students, physicians or other community groups with valid and reliable measures. We provide the first German measure for assessing this construct. The German AAS is a brief measure with good acceptance and satisfying psychometric properties. In a next step, validation in diverse settings and as a 13-item version without item 10 as well as assessment of convergent and predictive validity of the German AAS should be evaluated. The AAS has the potential to facilitate and enhance the assessment of attitudes towards abortion in Germany, potentially paving the way for targeted interventions aimed at reducing barriers and improving care for individuals facing unintended pregnancies.

## Data Availability

The data underlying the results presented in the study are available from the corresponding author.

## LIST OF ABBREVIATIONS

AAS: Abortion Attitude Scale
AIC: Akaike Information Criterion
AL: Anja Lindig
AMOS: Analysis of Moment Structure, Statistical Package for the Social Sciences, International Business Machines Corporation
BIC: Bayesian Information Criterion
CFA: Confirmatory Factor Analysis
Chi^2^: Discrepancy Chi-squared Statistic
CFI: Comparative Fit Index
COSMIN: Consensus-based Standards for the Selection of Health Measurement Instruments
Df: Degree of freedom
EC: Eva Christalle
JZ: Jördis Zill
KMO: Kaiser-Meyer-Olkin criterion
MB: Mirja Baumgart
MLR: maximum likelihood estimator
MR: Mareike Rutenkröger
RMSEA: Root Mean Square Error of Approximation
SPSS: Statistical Package for the Social Sciences, International Business Machines Corporation
SRMR: Standardized Root Mean Squared Residual
TLI: Tucker-Lewis Index
TRAPD: Translation, Review, Adjudication, Pretesting, Documentation

## DECLARATIONS

### Ethics approval and consent to participate

This study was carried out according to the latest version of the Helsinki Declaration of the World Medical Association, respected principles of good scientific practice and met standards of research ethics. This study was approved by the Local Ethics Committee of the University Medical Centre Hamburg-Eppendorf, Germany (LPEK-0628, May 27^th^, 2023). Data protection and confidentiality requirements were met. Study participation was voluntary. All participants received information about the aims of the study, data collection, and the use of collected data and gave informed consent before participation.

### Availability of data and materials

The dataset collected and analyzed during this study is available from the corresponding author on reasonable request.

The study was registered via AsPredicted.org prior to data collection (September 8^th^, 2023). Trial registration number: 142964

### Author contributions

AL and MB conceptualized, prepared and conducted this secondary analysis. MB and AL were involved in the translation and adaptation process. MB collected the data for psychometric evaluation. AL conducted the analysis in collaboration with EC. JZ was principal investigator of the CarePreg study and supervised the study at hand. All authors contributed to the interpretation of results. AL drafted the manuscript and MB, EC and JZ were involved in critically revising the manuscript for important intellectual content. All authors gave final approval of the version to be published.

## Acknowledgements

We thank all participants of the online survey. We thank our cooperation partners (counselors in social support service and physicians providing abortions) for their critical and important feedback during the development of the online survey. We also thank Maja Lina Böcher for her help preparing this manuscript. We especially thank Dr. Mareike Rutenkröger for supporting the team during the process of translation and adaptation.

## Checklist for reporting statement

To report the results of this validation study, we used the Authors’ Guidelines for Reporting Scale Development and Validation Results by Cabrera-Nguyen (see S3 Appendix). For assessment of comprehensibility as part of content validity, we used the COSMIN criteria (Consensus-based standards for the selection of health measurement instruments).

## Additional Files

S1 Appendix: Details on item translation and adaptation

S2 Appendix: Further demographic data and inter-item correlations

S3 Appendix: Checklist for reporting standards

